# Effects of the COVID-19 pandemic on sympathectomies in Brazil’s public health system

**DOI:** 10.1101/2024.05.13.24307276

**Authors:** Carolina Carvalho Jansen Sorbello, Felipe Soares Oliveira Portela, Giulia St. Sève Girardi, Marcelo Fiorelli Alexandrino da Silva, Marcelo Passos Teivelis, Nelson Wolosker

**Affiliations:** Hospital Israelita Albert Einstein, São Paulo, SP, Brasil; Universidade de São Paulo - USP, Faculdade de Medicina, Departamento de Cirurgia Vascular e Endovascular, São Paulo, SP, Brasil; Faculdade Israelita de Ciências da Saúde Albert Einstein, São Paulo, SP, Brasil

**Keywords:** Big data, hyperhidrosis, endoscopic thoracic surgery, sympathectomy, public health, COVID-19

## Abstract

2.

**Objective:** To analyze and compare the number of sympathectomies for the treatment of hyperhidrosis in the public health system (SUS - Unified Health System) before, during, and after the peak of the COVID-19 pandemic in Brazil.

**Method:** A retrospective cross-sectional study, in accordance with the guidelines of the Research Ethics Committee, which analyzed the number of elective surgical procedures performed in SUS-Brazil, including the number of thoracic sympathectomies for the treatment of hyperhidrosis, in the 2 years prior to the COVID-19 pandemic (2018 and 2019) as well as in the 2 years of maximum prevalence (2020-2021) and in the 2 years following the peak of cases (2022-2023). The data analyzed was obtained from the database of the Department of Informatics of the Unified Health System (DATASUS).

**Results:** The number of thoracic sympathectomies performed in the SUS during the years 2020 and 2021 significantly decreased, probably secondary to the allocation of resources to the care of COVID-19 patients. This was followed by a lower than expected increase in the number of sympathectomies compared to pre-pandemic data.

**Conclusion:** The volume of sympathectomies, and consequently the surgical treatment of hyperhidrosis patients dependent on the SUS, have been extremely affected by the COVID-19 pandemic and remain in slow recovery, with no short-term prospect of returning to previous figures.

## 3. Introduction

Hyperhidrosis is a condition that seriously compromises the personal life ^1^, professional, and mental well-being ^2^ of individuals who experience it. Most people with this condition are teenagers or young adults who are at the peak of their productive lives.^3^ When surgically treated with sympathectomy with adequate indication, the risk is low ^4^ and the procedure improves quality of life in over 90% of cases.^5^

In Brazil, approximately 75% of the population, which is around 170 million people, depend on the public system (SUS) ^6^ to meet their health needs. Data regarding public health in Brazil, including information about surgical procedures carried out by the SUS (Unified Health System), is stored and managed on a single portal belonging to the Department of Information and Informatics of the Unified Health System (DATASUS). ^7^ The Digital Health and Information Secretariat manages this system, which stores data on health indicators, health care, epidemiological and morbidity information, information on the health care network, vital statistics, demographic and socio-economic information, financial information, and public health costs. The information is anonymized and available on the DATASUS online platform, which is the basis for data collection in this study.

Louzada et al ^8^, conducted a nationwide cross-sectional study that analyzed 12 years of data about thoracic Sympathectomy in Brazil.^9^ They found that from 2008 to 2019, 13,201 endoscopic thoracic sympathectomies were performed in the country, with a rate of 68.44 procedures per 10 million inhabitants per year. Another population-based cross-sectional study by Da Silva et al.^10^, showed that the number of sympathectomies decreased significantly over 11 years (P = 0.001), probably due to the increased use of Oxybutynin as a drug treatment. ^11,12^

In the 2020 and 2021, the country became one of the global epicenters of the COVID-19 pandemic ^13^, with over 38 million cases and more than 700.000 deaths associated with the disease.^14^ The healthcare system had to be reorganized to deal with respiratory syndrome cases ^15^ and elective surgical treatments, such as sympathectomies, were unavoidably postponed. This caused a backlog of patients and further impacted the quality of life of these individuals.

Although some previous studies have evaluated the negative impact of elective surgeries during the peak of the COVID-19 pandemic, there is a gap in our understanding. No study has yet evaluated the trend in the number of sympathectomies after the end of the COVID-19 pandemic.^16^ This underscores the need for further research to fully comprehend the long-term effects of the pandemic on our healthcare system.

This study used DATASUS data to analyze trends in the number of sympathectomies in Brazil’s public health system in the pre-pandemic years (2018 and 2019), compared to the period during the pandemic (2020 and 2021) and after (2022-2023) the COVID-19 outbreak.

## 4. Methods

This was a study where we analyzed data obtained from DATASUS, a Ministry of Health platform that gathers information on hospitalizations financed by SUS. The study was approved by the institution’s Research Ethics Committee. This platform is public and is fed with unidentified data, which is why there was no need to apply an informed consent form.

### Period analyzed, data source and extraction

This retrospective study analyzed the available data on thoracic sympathectomies performed in Brazil’s public health network between 2018 and 2023. The data was obtained from DATASUS, a digital platform of the Unified Health System that offers open data on procedures carried out in public hospitals accredited by the system. This accreditation is mandatory for the government to reimburse institutions.

All the data was collected on this platform using an automated extraction method programmed by the institution’s IT service in the Python language (v. 2.7.13; Beaverton, OR, USA), using the Windows 10 operating system. Field selection on the DATASUS platform and subsequent table adjustment were carried out using Selenium WebDriver (v. 3.1.8; Selenium HQ) and Pandas (v. 2.7.13; Lambda Foundry, Inc. and PyData Development Team, NY, USA).

### Selection of thoracic sympathectomy procedures on the platform

Data was selected and collected for the following coded procedure, according to the Brazilian public health system’s coding: 0403050146 video-assisted thoracic sympathectomy.

For the procedure selected, demographic data (distribution of sympathectomies by region of the country, age group, and gender of patients), as well as data on length of stay and reimbursement amounts passed on to institutions for the surgeries performed, were collected.

The data was compiled in .cvs format and organized into tables using Microsoft Office Excel 2016 (Redmond, WA, USA). The country’s population data by age group was obtained from the website of the Brazilian Institute of Geography and Statistics (IBGE). Calculations of the standardized rates were based on the population exclusively dependent on the Unified Health System (SUS) services, published periodically by the National Health Agency (ANS), based on data released in December of each year.

### Ethics Committee approval and statistical analysis

The data is obtained anonymously through this platform, so a consent form was unnecessary. The project was approved by the institution’s ethics committee.

For this study, we initially analyzed the number of video-assisted thoracic sympathectomies performed in the SUS from 2018 to 2023, along with the number of sympathectomies divided by macro-region, age, gender and race in the same period. The length of stay, costs, and clinical-surgical outcomes of patients undergoing sympathectomy in these 6 years were also analyzed. Additionally, the volume of sympathectomies in 3 periods: before the COVID-19 pandemic (2018-2019), during the height of the pandemic (2020-2021), and the outbreak of COVID-19, was comparatively analyzed.

The statistical analysis was conducted using SPSS 20.0 for Windows (IBM Corp, Armonk, NY). Linear regression was employed to analyze the evolution of sympathectomy rates by age group. The chi-square and likelihood ratio tests were used to compare thoracic sympathectomy rates by macro-region of the country and mortality rates by region. For all tests, a value of P ≤ 0.05 was considered statistically significant.

## 5. Results

Figure 1 depicts the temporal analysis of the number of sympathectomies. Prior to the pandemic (2018 and 2019), an average of 863 procedures were carried out every year. However, during the COVID-19 pandemic, the average number of procedures fell to approximately 347 per year, which is a significant reduction of nearly 60% compared to the earlier period. After the pandemic, with the full resumption of elective surgeries, there was a trend towards an increase in sympathectomies, but it was still lower than the pre-pandemic levels. In 2023, there were around 30% fewer sympathectomies performed than in 2018/19.

**Figure 1:**
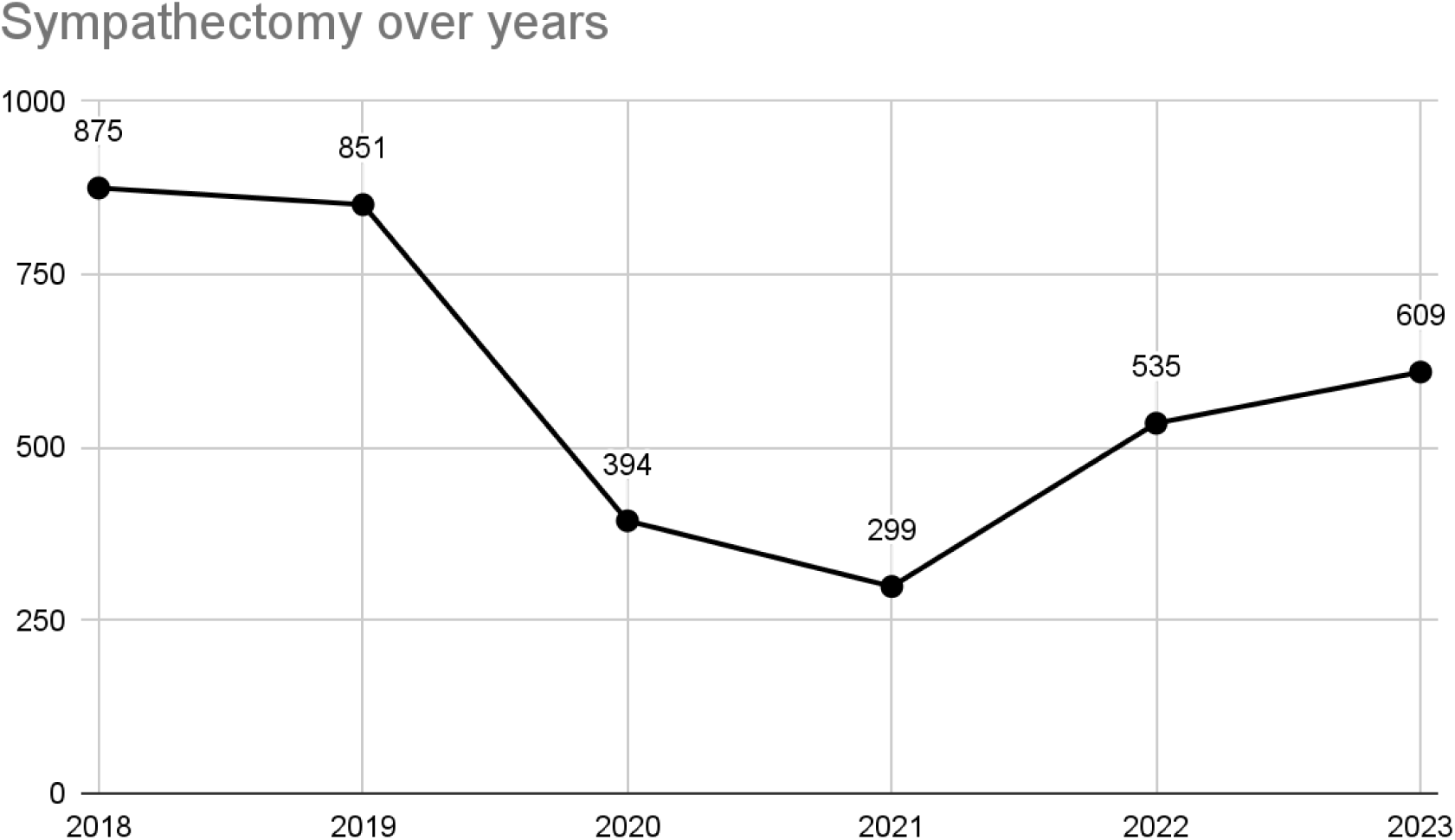
Temporal trend in the number of sympathectomies in Brazil in 3 periods: pre-pandemic (2018 and 2019), during the pandemic (2020 and 2021) and post-pandemic (2022-2023).

Figure 2 presents an analysis of the number of sympathectomies performed per macro-region. The data shows that the majority of the procedures took place in the Southeast region, accounting for almost 50% of all surgeries in the country. The South region came in second place with over 30% of the sample between 2018 and 2023.

**Figure 2:**
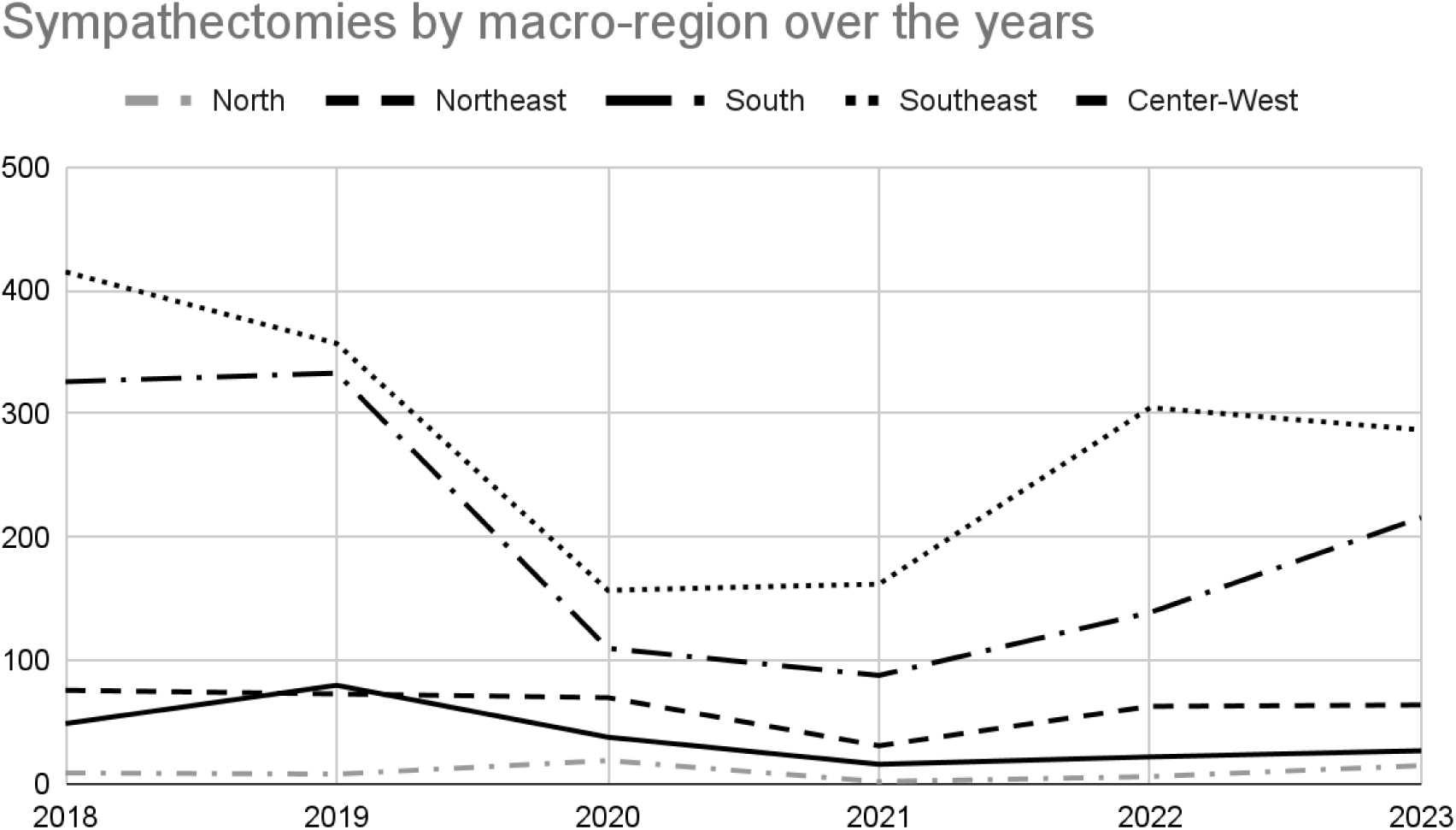
Temporal trend in the number of sympathectomies in Brazil, divided by macro-region, in 3 periods: pre-pandemic (2018 and 2019), during the pandemic (2020 and 2021) and post-pandemic (2022-2023) (North, Northeast, South, Southeast and Center-West).

It is important to note that the pandemic (2020-2021) had a negative impact on surgical volume, and this trend was observed in all regions, except the North. However, the Northeast and Midwest regions showed less evidence of downward trend.

According to the analysis presented in Figures 3 and 4, there were no significant differences in the patient profile between the periods studied, in terms of gender and age. In all cases, more than 80% of the patients who underwent surgery were between 15-59, and women were more prevalent, accounting for approximately 65% of the sympathectomies performed during the analyzed periods.

**Figure 3:**
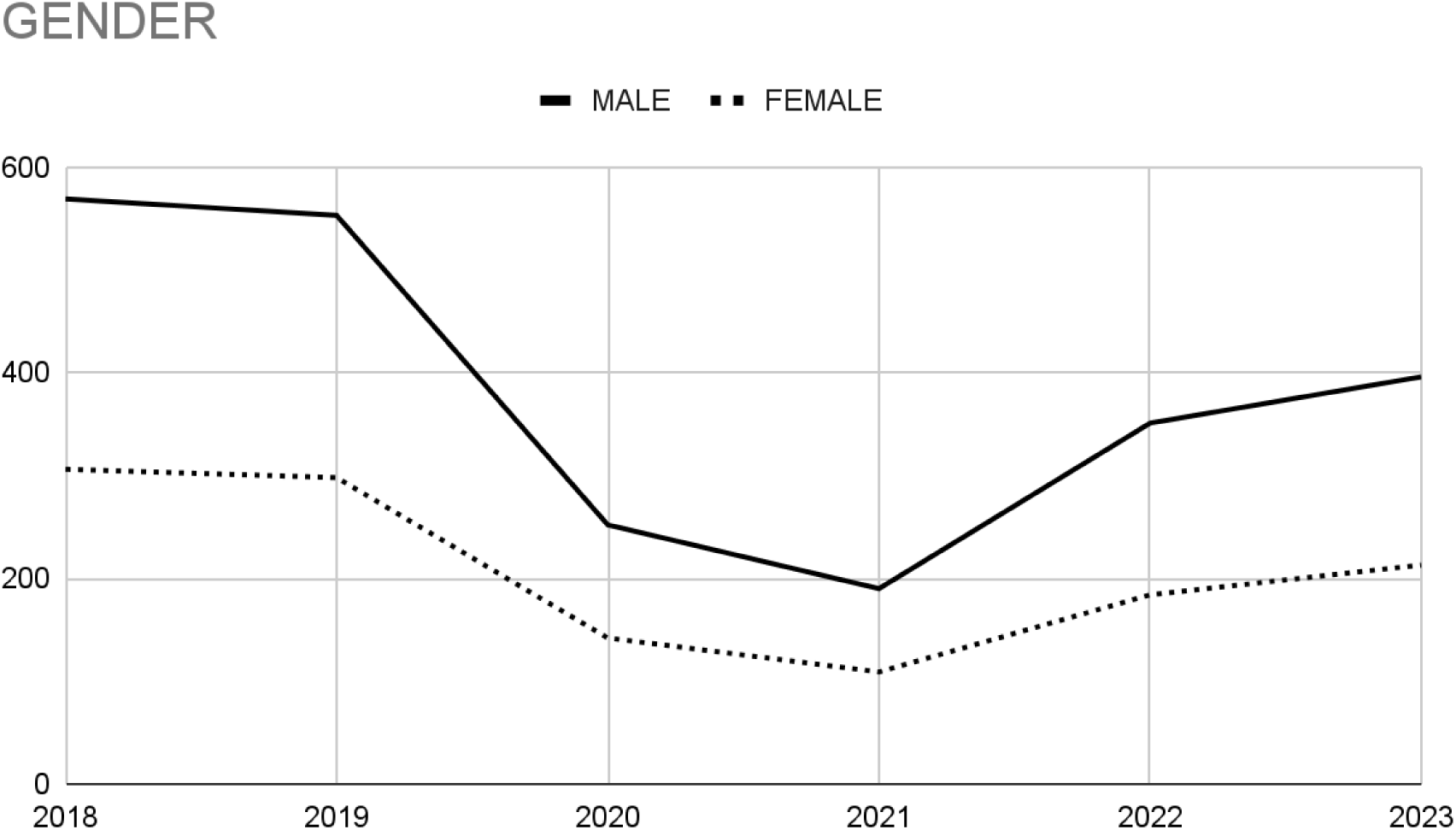
Temporal analysis of the gender profile of patients undergoing sympathectomy from 2018 to 2023.

**Figure 4:**
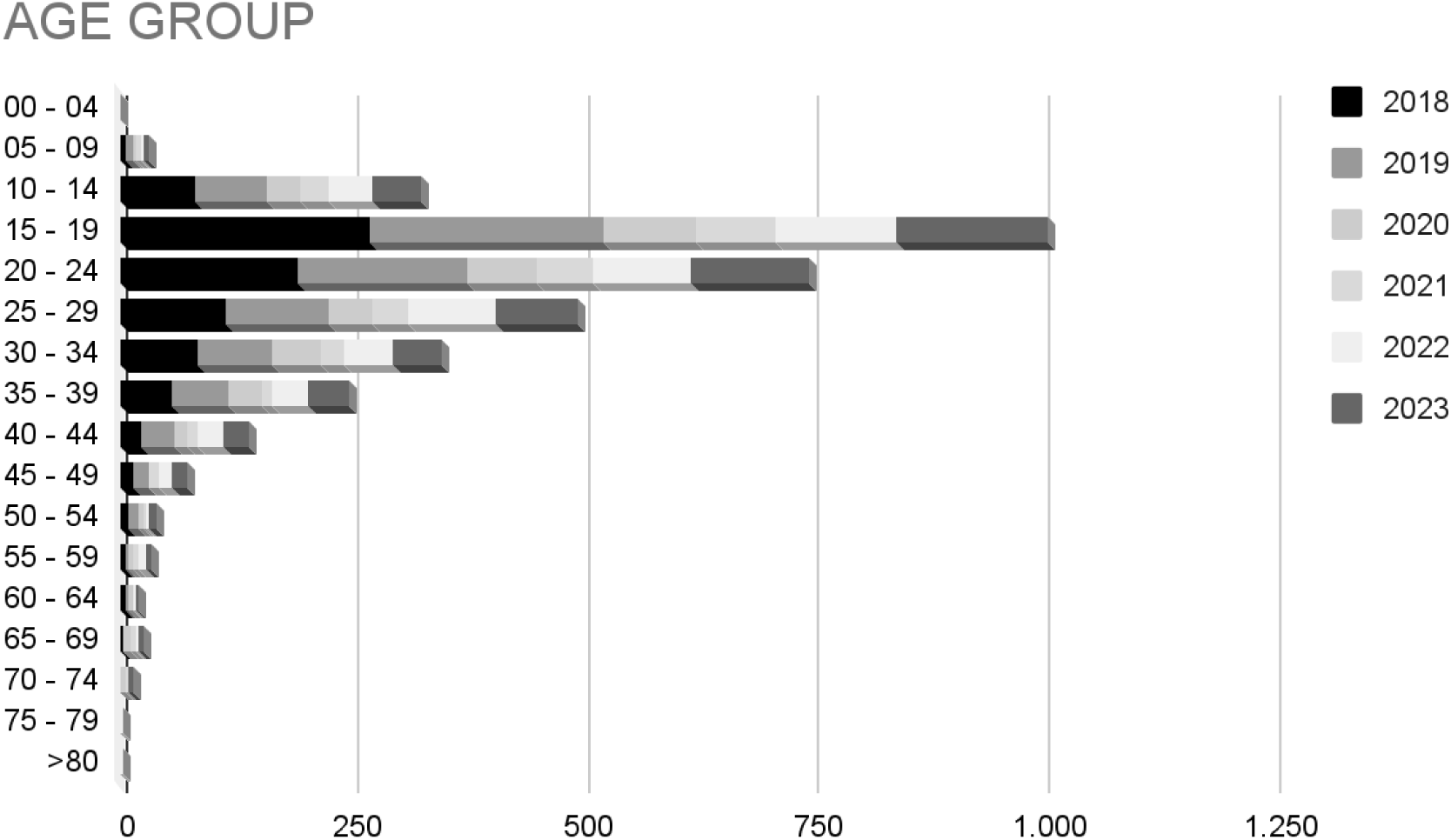
Temporal analysis of the age of patients undergoing sympathectomy from 2018 to 2023.

In Table 1 and Figure 5, the clinical-surgical aspects related to sympathectomy are presented. The analysis shows that the length of hospital stays was less than 3 days in 90% of cases throughout the analyzed period. Additionally, the cost per surgery remained stable over the years, staying below 1.500,00 reais.

**Figure 5:**
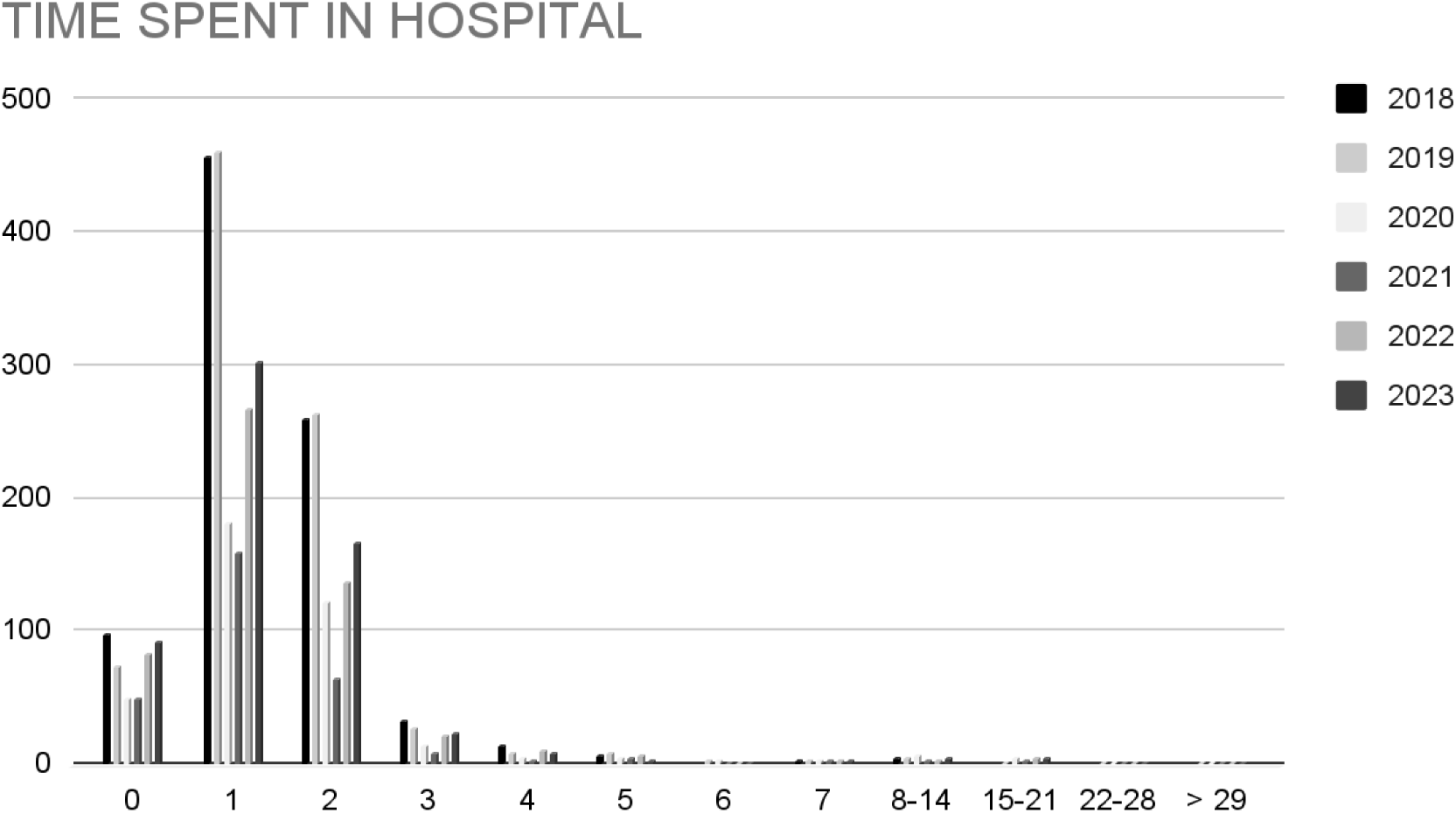
Temporal analysis of hospital stay related to sympathectomy, from 2018 to 2023.

**Table 1:** Temporal analysis of costs per procedure, in each macro-region, related to sympathectomy, from 2018 to 2023.

| Year | 2018 | 2019 | 2020 | 2021 | 2022 | 2023 |
| --- | --- | --- | --- | --- | --- | --- |
| Region |  |  |  |  |  |  |
| North | R\$1.028,98 | R\$1.030,15 | R\$1.029,86 | R\$1.032,52 | R\$1.039,33 | R\$1.076,46 |
| Northeast | R\$1.067,28 | R\$1.061,55 | R\$1.133,65 | R\$1.190,95 | R\$1.107,85 | R\$1.134,70 |
| South | R\$1.031,01 | R\$1.050,14 | R\$1.102,88 | R\$1.033,21 | R\$1.136,58 | R\$1.058,39 |
| Southeast | R\$1.055,68 | R\$1.059,86 | R\$1.369,95 | R\$1.220,46 | R\$1.120,67 | R\$1.115,30 |
| Midwest | R\$1.126,61 | R\$1.104,86 | R\$1.192,07 | R\$1.311,27 | R\$1.068,63 | R\$1.144,75 |

During the peak of the pandemic, the need for ICUs in relation to sympathectomy increased only in the Southeast. On the other hand, in the other regions, the number of patients referred to the ICU remained stable. However, there was no significant difference in the number of deaths over the six years.

## 6. Discussion

The COVID-19 pandemic has caused significant changes to health services across the globe ^17^, as resources have been redirected to care for patients affected by the coronavirus. Brazil is no exception to this, as emergency care has been prioritized, with elective surgeries and minor medical care receiving less attention.^18^

During the COVID-19 pandemic, medical societies, including the American College of Surgeons, classified surgical procedures into different groups. They recommended suspending elective surgeries ^19^, especially those for diseases that have lower potential for morbidity and mortality, like sympathectomies for hyperhidrosis.^20^ As a result, "purely elective" surgeries were initially canceled in nearly all specialties, leading to the suspension of over 900,000 elective surgeries in Brazil in 2020 alone.^21^ Some of these surgeries have started to resume by the end of 2021.

The postponement of non-urgent medical procedures during the pandemic has had consequences for both patients and the healthcare system. Patients suffered prolonged delays, which affected their quality of life, productivity and, in some cases, culminated in the progression of the disease. For the health system, the backlog of cases means increased spending in the post-pandemic period to reduce the waiting list for surgeries.

Although the number of sympathectomies has slowly returned to pre-pandemic levels, it is still significantly lower than the average in the pre-pandemic period. There are several possible reasons for this, including the economic crisis resulting from the pandemic, the need to prioritize more urgent procedures, loss of follow-up, and limited access to health services.

The number of sympathectomies varies by region, with more developed regions recovering more quickly than less developed regions in the post-pandemic period. Thus, states in the South and Southeast experienced a faster and more effective economic recovery in the post-pandemic period, facilitating reinvestment in elective surgeries.

The demographic profile of patients undergoing sympathectomies has remained stable before, during, and after the pandemic. There was a peak incidence in young patients aged between 15 to 30 years, with a majority of females.^22,23^ This trend is consistent with previous studies and suggests that women are more health-conscious than men. Young patients, who suffer from hyperhidrosis and have a lower surgical risk, have a significant social impact and, therefore, exhibit a greater demand for treatment.^24^

This study reaffirms the previously established facts regarding sympathectomies. The surgery has low morbidity and mortality rates and requires only a short stay in the hospital.^25^ Ideally, patients should be admitted the day prior to the procedure and discharged on the first day after the surgery, following a brief stay in the ward. This will reduce unnecessary hospital occupancy and avoidable costs. In this context, sympathectomy is a low-cost surgery ^26^, with a cost of approximately 300 dollars per procedure, with little change in values during the pandemic despite the shortage of essential supplies.

Due to the COVID-19 outbreaks, several surgeries of varying complexity have been suspended to prioritize the treatment of patients with the potential to experience severe morbidity. This has resulted in significant delays for many procedures. As we return to normal, surgeries for more severe diseases such as cancer, transplants, and heart surgery, will be prioritized over procedures like sympathectomies. Despite the considerable impact on the quality of life of patients ^27^, hyperhidrosis is not typically life-threatening ^28^ and will likely take longer to return to pre-pandemic levels than other elective procedures.

## 7. Conclusion

The COVID-19 pandemic has had a severe impact on the number of sympathectomies being performed. Although there has been an upward trend in the number of this kind of surgery, it is unlikely that we will see a return to pre-pandemic levels anytime soon. However, the pandemic has not significantly affected the demographic profile, costs per procedure, length of hospital stay, and mortality associated with sympathectomy.

## Data Availability

All data produced in the present work are contained in the manuscript All data analyzed are available online at https://datasus.saude.gov.br/

---

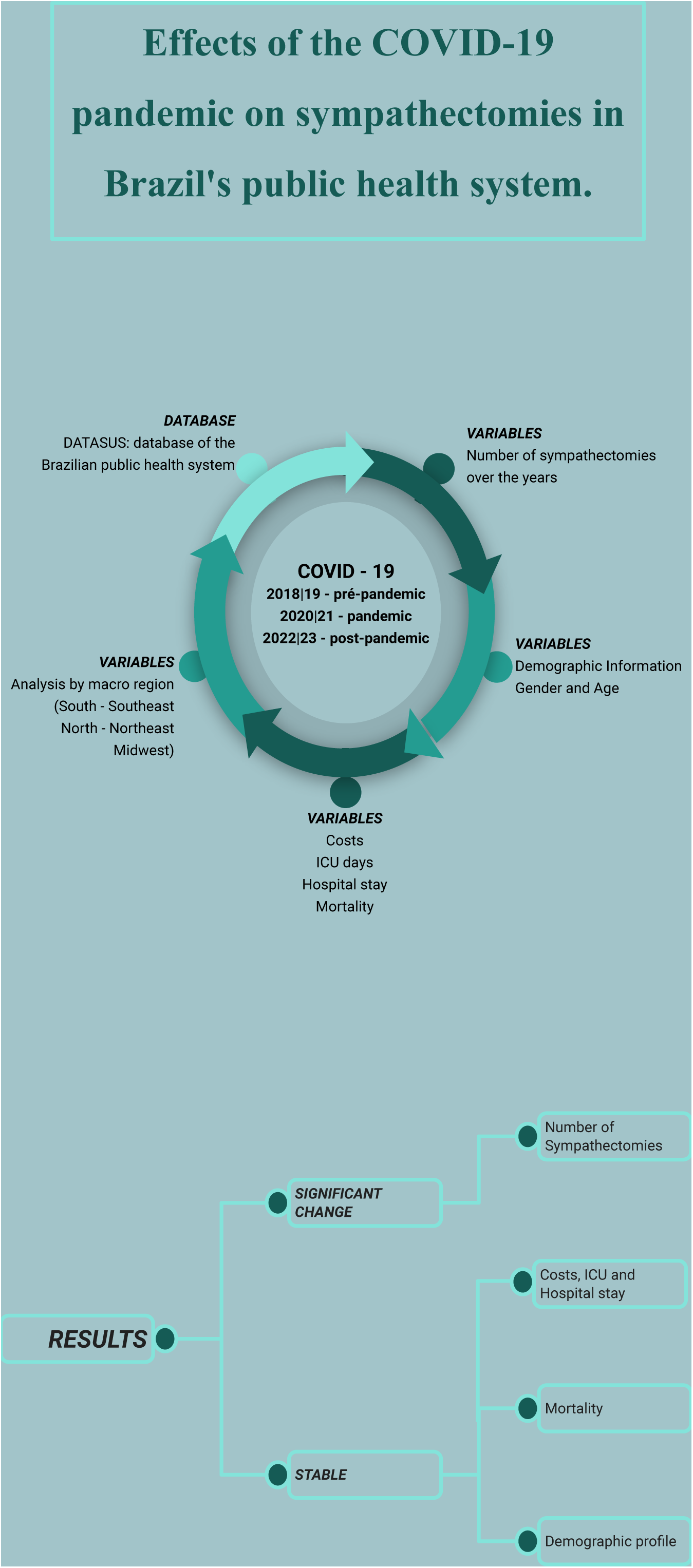

